# Waist-to-Height Ratio Identifies Cardiometabolic Risk Not Captured by Body Mass Index: The All of Us Research Program

**DOI:** 10.64898/2026.11.04.26367325

**Authors:** Adam W. Potter, Caroline E. Murphy, Jason H. Wasfy, Karl E. Friedl

**Author notes:** Corresponding author: Adam W. Potter, 10 General Greene Ave., Natick, MA 01760.

## Abstract

**Background:** Body mass index (BMI) does not characterize central adiposity and may obscure cardiometabolic risk among adults classified as normal weight. We evaluated associations between waist-to-height ratio (WHtR) and incident disease across the BMI spectrum and among adults with normal BMI.

**Methods:** We studied 308,600 adults in the All of Us Research Program. Participants with the corresponding prevalent disease were excluded separately for eight cardiometabolic outcomes. Cause-specific Cox models adjusted for demographic, socioeconomic, smoking, and BMI-related factors. We evaluated continuous and nonlinear WHtR associations, elevated WHtR among normal-BMI adults, and incremental discrimination. Additional analyses tested WHtR-by-BMI-category interactions.

**Results:** Among 77,684 adults with normal BMI, 24,301 (31.3%) had WHtR ≥0.50. In whole-cohort analyses, adjusted hazard ratios (HRs) per 0.05 higher WHtR ranged from 1.07 to 1.31 across the eight outcomes. In BMI-stratified interaction models, corresponding diabetes HRs were 1.52 (95% CI, 1.43–1.61) in normal BMI, 1.42 (1.36–1.49) in overweight, and 1.22 (1.20–1.25) in obesity (P for interaction <0.001). Hypertension associations also differed across BMI groups (P for interaction <0.001). Among normal-BMI adults, WHtR ≥0.50 versus <0.50 was associated with diabetes (HR, 1.74), myocardial infarction (1.49), and ischemic stroke (1.52). Associations with all eight outcomes persisted in normal-BMI landmark analyses excluding the first year. Flexible age adjustment preserved the principal findings. Adding nonlinear WHtR improved cross-validated discrimination, most notably for diabetes (ΔC-index=0.0134).

**Conclusions:** WHtR identified cardiometabolic risk beyond BMI classification. Associations with diabetes and hypertension were particularly pronounced among normal-BMI adults, supporting assessment of central adiposity alongside BMI.

**Clinical Perspective:** *What Is New?:*

- In this longitudinal cohort of 308,600 adults, higher waist-to-height ratio (WHtR) was associated with incident type 2 diabetes, hypertension, ischemic heart disease, myocardial infarction, ischemic stroke, heart failure, atrial fibrillation, and chronic kidney disease independently of body mass index (BMI).
- Among adults with normal BMI, 31.3% had WHtR ≥0.50, identifying a phenotype associated with higher incidence of all eight evaluated cardiometabolic outcomes. These associations persisted after excluding the first year of follow-up.
- The association between continuous WHtR and incident diabetes was stronger among adults with normal BMI than among those with obesity. For hypertension, the association was stronger in normal-BMI adults than in both overweight and obesity.

*What Are the Clinical Implications?:*

- BMI alone can miss clinically relevant central adiposity and associated cardiometabolic risk among adults classified as having normal weight.
- WHtR is a practical complementary measure that clinicians may consider alongside BMI during preventive cardiometabolic assessment, particularly when a patient has a normal BMI but other risk factors or clinical concerns.
- WHtR should not be treated as a stand-alone risk model or a universal treatment threshold; its incremental prognostic value and exposure-response patterns differed by outcome.

## Introduction

Body mass index (BMI) provides a practical framework for surveillance of overweight and obesity but does not distinguish fat from lean tissue or characterize fat distribution. Individuals with the same BMI can therefore differ substantially in abdominal adiposity and cardiometabolic risk, including among adults classified as normal weight (Ross et al., 2020; Powell-Wiley et al., 2021; Potter et al., 2025; Potter and Friedl, 2025).

BMI’s limitations vary across populations (Hubbard, 2000). Asian populations may develop greater body fatness and cardiometabolic risk at traditionally normal BMI values (Deurenberg-Yap et al., 2000; Deurenberg et al., 2002). A Lancet Diabetes & Endocrinology Commission proposed supplementing BMI with confirmation of excess adiposity and assessment of organ dysfunction or functional limitations (Rubino et al., 2025). Waist measurement, incorporated into Japan’s national screening program, partly addresses these limitations, but fixed waist thresholds remain sensitive to stature and body habitus (Hirata & Uchida, 2026).

Waist-to-height ratio (WHtR) scales waist circumference to stature. Meta-analyses and prospective studies suggest that WHtR identifies cardiometabolic risk at least as well as BMI and other anthropometric measures (Ashwell et al., 2009; Ashwell et al., 2012; Savva et al., 2013; Gelber et al., 2008; Jayedi et al., 2022; Feng et al., 2024). The UK National Institute for Health and Care Excellence, recommends WHtR alongside BMI <35 kg/m^2^, identifying 0.50 as a practical boundary for increased central adiposity (NICE, 2026). An important question remains unresolved: does WHtR identify prospective disease risk that is meaningfully missed when BMI is used to classify apparently normal weight adults?

Most WHtR studies address prevalent risk factors, individual outcomes, or mortality. Although waist circumference and waist-to-hip ratio identify cardiovascular risk heterogeneity within BMI categories, comparable longitudinal evidence for WHtR across multiple outcomes remains limited (Dardari et al., 2026). Using data from the All of Us Research Program (AoURP), we evaluated WHtR associations with eight incident cardiometabolic outcomes, emphasizing risk among normal-BMI adults, nonlinear relationships, standardized event probabilities, and incremental discrimination beyond BMI (AoURP Investigators, 2019).

## Methods

### Study Design and Data Source

We conducted a longitudinal study using AoURP Registered Tier Curated Data Repository version 9 (R2025Q4R6; cutoff January 1, 2025), which integrates physical measurements, surveys, and EHRs standardized to the Observational Medical Outcomes Partnership Common Data Model (OMOP). Analyses used the secure Researcher Workbench; no participant-level data were exported. The U.S. Army Research Institute of Environmental Medicine Office of Research Quality and Compliance determined that the analysis met exemption criteria (Protocol #27-01R).

### Study Population and Anthropometric Measures

The index date was the first numeric AoURP-computed waist measurement. Eligible adults were aged ≥18 years, had valid same-day waist and height measurements, met anthropometric quality-control criteria, and had ≥365 days of pre-index observation to ascertain prevalent disease. The longitudinal cohort included 308,600 participants. Sex-adjusted analyses included participants recorded as female or male; other categories had insufficient numbers for stable outcome-specific estimation. Each outcome-specific risk set excluded the corresponding prevalent disease on or before baseline.

BMI–WHtR phenotypes could be assigned to 308,218 participants; 382 lacked valid analytic BMI (**Supplemental Figure S1; Supplemental Table S1**). Program-computed waist circumference is the mean of the two closest measurements. WHtR was waist circumference divided by same-day height, both in centimeters. BMI used the program-computed measure; BMI calculated from same-day height and weight showed near-perfect agreement.

Prespecified cohort-entry anthropometric quality-control limits were 120 to 220 cm height and 40 to 200 cm waist circumference. BMI values from 10 to 80 kg/m^2^ were considered valid for BMI-based analyses; BMI validity was assessed separately from eligibility for the longitudinal anthropometric cohort. BMI was categorized as underweight (<18.5 kg/m^2^), normal weight (18.5 to <25 kg/m^2^), overweight (25 to <30 kg/m^2^), or obesity (≥30 kg/m^2^). For BMI–WHtR discordance analyses, elevated WHtR was defined a priori as ≥0.50. Thresholds of 0.52 and 0.55 were evaluated in sensitivity analyses and were not selected through outcome-specific optimization.

### Cardiometabolic Outcomes

We evaluated eight incident outcomes: type 2 diabetes (T2D), hypertension, ischemic heart disease (IHD), myocardial infarction (MI), ischemic stroke, heart failure (HF), atrial fibrillation (AF), and chronic kidney disease (CKD). Outcomes were constructed from standardized OMOP condition concepts and clinically relevant descendants. Concept hierarchies were adjudicated before outcome construction to distinguish concepts suitable for identifying prevalent disease from those suitable for initiating an incident event.

Pregnancy-related conditions, historical or status concepts, procedural diagnoses, and other concepts not representing new-onset clinical disease were excluded from incident definitions, as appropriate. For each outcome, participants with corresponding prevalent disease on or before the index date were excluded. An incident event was defined as the first qualifying diagnosis after the index date (**Supplemental Table S2**).

### Follow-Up and Death Censoring

Follow-up extended from baseline to the first qualifying outcome, observation end, or death, whichever occurred first. Events recorded on the date of death were retained because within-day temporal ordering could not be established and terminal diagnoses may represent legitimate clinical events. Diagnoses recorded after death were excluded. Primary models therefore estimated cause-specific hazards for each incident cardiometabolic outcome. Within each fully adjusted analytic population, follow-up was summarized as median (IQR), and person-time was summed from baseline to the first event or censoring.

### Covariates

Models adjusted for baseline age, sex, continuous BMI, race, ethnicity, smoking (never, former, current), education (<high school, high school, vocational training, college graduate/advanced degree), and household income using available AoURP categories.

### Statistical Analysis

Analyses used R in the Researcher Workbench. Baseline characteristics were summarized overall and by BMI–WHtR phenotype as mean (SD) or n (%), without between-group hypothesis tests. Whole-cohort analyses evaluated continuous/nonlinear WHtR associations and incremental discrimination in outcome-specific complete-case risk sets. Normal-BMI analyses evaluated WHtR ≥0.50 versus <0.50 and standardized event probabilities.

Primary inferential analyses used cause-specific Cox proportional hazards regression. WHtR was initially modeled continuously, with hazard ratios (HRs) reported per 0.05-unit higher WHtR; estimates per 1-SD higher WHtR were also calculated. Nonlinear associations were evaluated using restricted cubic splines with 4 knots placed at approximately the 5th, 35th, 65th, and 95th percentiles of the WHtR distribution. WHtR 0.50 was used as the graphical reference value. Likelihood-ratio tests compared nonlinear with linear model specifications.

To evaluate discordance between BMI and central adiposity, analyses were restricted to adults with normal BMI (18.5 to <25 kg/m^2^). WHtR ≥0.50 was compared with WHtR <0.50. Continuous BMI was retained to reduce residual confounding by body size within the normal-BMI range.

Additional exploratory analyses evaluated whether continuous WHtR associations differed across normal-BMI, overweight, and obesity categories. Participants with underweight BMI were excluded. Cause-specific Cox models included BMI-category-specific baseline hazards, WHtR expressed per 0.05-unit increment, and WHtR-by-BMI-category interaction terms. Models retained continuous BMI and the other adjustment covariates, whose coefficients were shared across BMI categories. BMI-specific WHtR estimates and pairwise ratios of hazard ratios were derived from the same interaction model. Two-degree-of-freedom likelihood-ratio tests compared models with and without the interaction terms. Benjamini–Hochberg adjustment was applied across the eight outcome-specific omnibus interaction tests separately within each age specification. Pairwise comparisons used two-sided Wald tests and were interpreted as exploratory without multiplicity adjustment (**Supplemental Tables S3 and S4**).

For exploratory nonlinear interaction analyses, WHtR was represented using a natural cubic spline with internal knots at the outcome-specific pooled 35th and 65th percentiles and boundary knots at the 5th and 95th percentiles. Six-degree-of-freedom likelihood-ratio tests compared models with a common WHtR spline association against models allowing BMI-category-specific associations. Benjamini–Hochberg adjustment was applied separately across the eight outcome-specific spline interaction tests within each age specification. Sensitivity analyses represented age using a natural cubic spline with 4 degrees of freedom (**Supplemental Tables S3 and S5**).

### Incremental Prognostic Information Beyond BMI

Nested Cox models compared the covariate model, including BMI, against models adding linear or spline WHtR. Fixed participant-level 5-fold cross-validation retained identical fold assignments across outcomes. Spline parameters were estimated within training sets to prevent leakage. Harrell’s C-index was calculated in held-out participants, with changes interpreted by magnitude and consistency (**Supplemental Table S6**).

### Standardized 3-Year Event Probabilities

Among normal-BMI adults, standardized 3-year event probabilities were estimated by predicting each participant’s probability under WHtR <0.50 and ≥0.50, retaining observed covariates, then averaging predictions. These cause-specific Cox estimates are model-based probabilities, not competing-risk cumulative-incidence estimates or causal contrasts.

## Sensitivity Analyses

Primary fully adjusted analyses used complete covariate data. Missing baseline covariates were addressed using multiple imputation with 20 imputations as a sensitivity analysis. Additional analyses evaluated WHtR thresholds of 0.50, 0.52, and 0.55.

A 365-day landmark analysis excluded participants who experienced the outcome or were censored during the first year after baseline to reduce potential influence from occult prevalent disease or near-term diagnostic ascertainment. Follow-up for retained participants was measured from day 365. Landmark analyses evaluated continuous WHtR in the whole cohort and WHtR ≥0.50 versus <0.50 among adults with normal BMI. Proportional-hazards assumptions were assessed using Schoenfeld residuals. As WHtR-specific nonproportionality was observed for hypertension and AF, interval-specific models estimated associations during 0 to 2 years and >2 years of follow-up. All hypothesis tests were 2-sided. Given the related outcomes and emphasis on estimation, findings were interpreted according to magnitude, precision, consistency, and robustness rather than binary statistical significance alone.

Age sensitivity analyses replaced linear age with a natural cubic spline (4 degrees of freedom), retaining the same complete-case populations for continuous WHtR and the normal-BMI WHtR ≥0.50 contrast.

Likelihood-ratio tests assessed sex-by-WHtR interactions, with sex-specific estimates from separate adjusted models, for continuous WHtR and the normal-BMI threshold contrast. Exploratory omnibus tests assessed race/ethnicity interactions in categories with adequate participants and events. Interpretation considered multiplicity, statistical power, and the social and contextual meaning of these classifications.

## Results

### Study Population and BMI–WHtR Phenotypes

The final longitudinal anthropometric cohort included 308,600 adults; 305,341 participants recorded as female or male were eligible for sex-adjusted inferential analyses. Before outcome-specific exclusions for prevalent disease, 207,346 participants had complete data for all fully adjusted covariates. Among 308,218 participants with sufficient anthropometric data for BMI–WHtR phenotype classification, 77,684 had normal BMI: 53,383 had WHtR <0.50 and 24,301 had WHtR ≥0.50. Thus, 31.3% of adults with normal BMI had WHtR ≥0.50. Among participants with normal BMI, those with WHtR ≥0.50 were older than those with WHtR <0.50 (mean age, 59.1 versus 47.0 years) and had greater waist circumference (89.2 versus 76.1 cm), despite a smaller difference in BMI (23.4 versus 22.0 kg/m²) (**Table 1; Supplemental Table S1**).

**Table 1.**
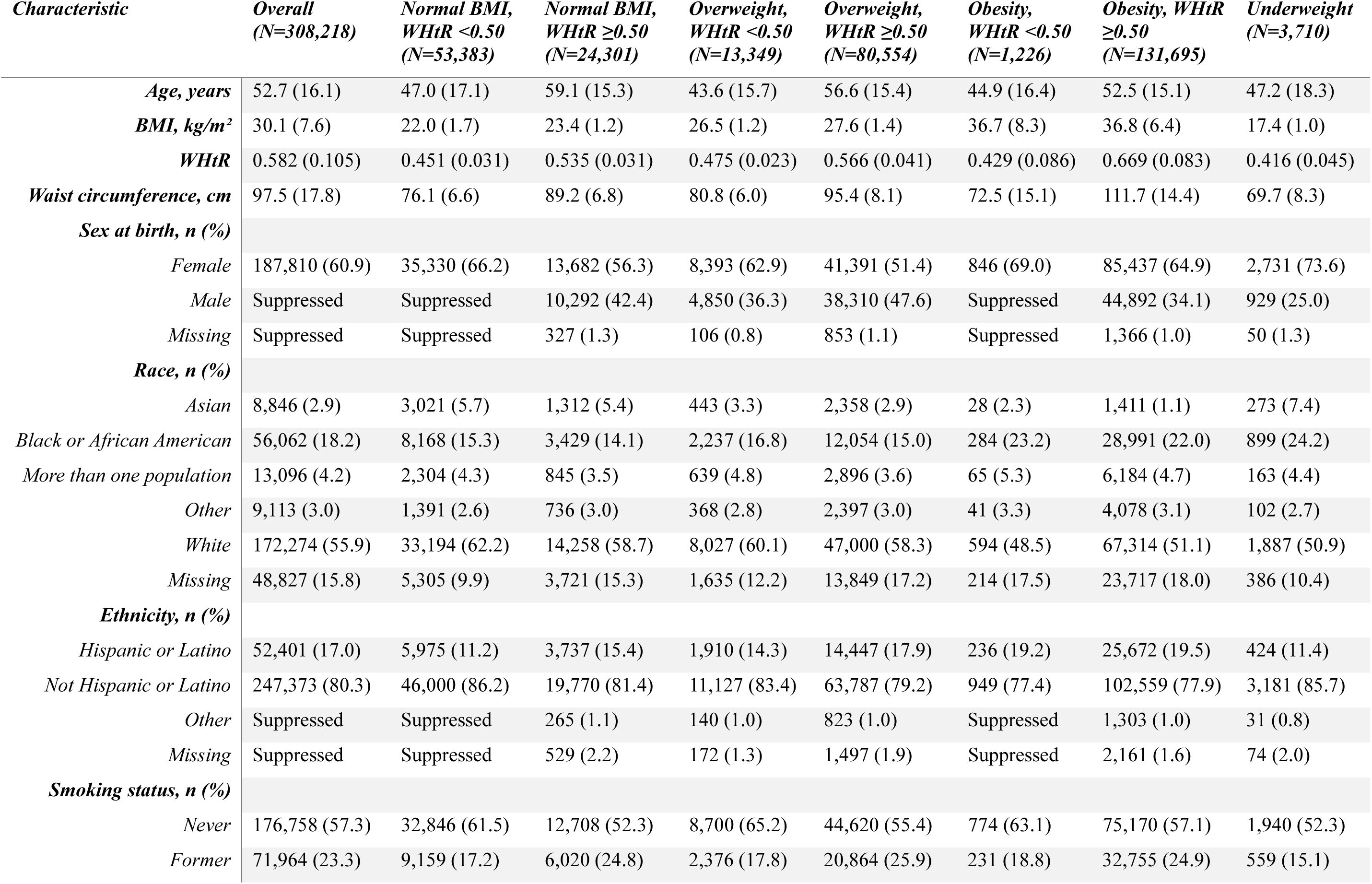
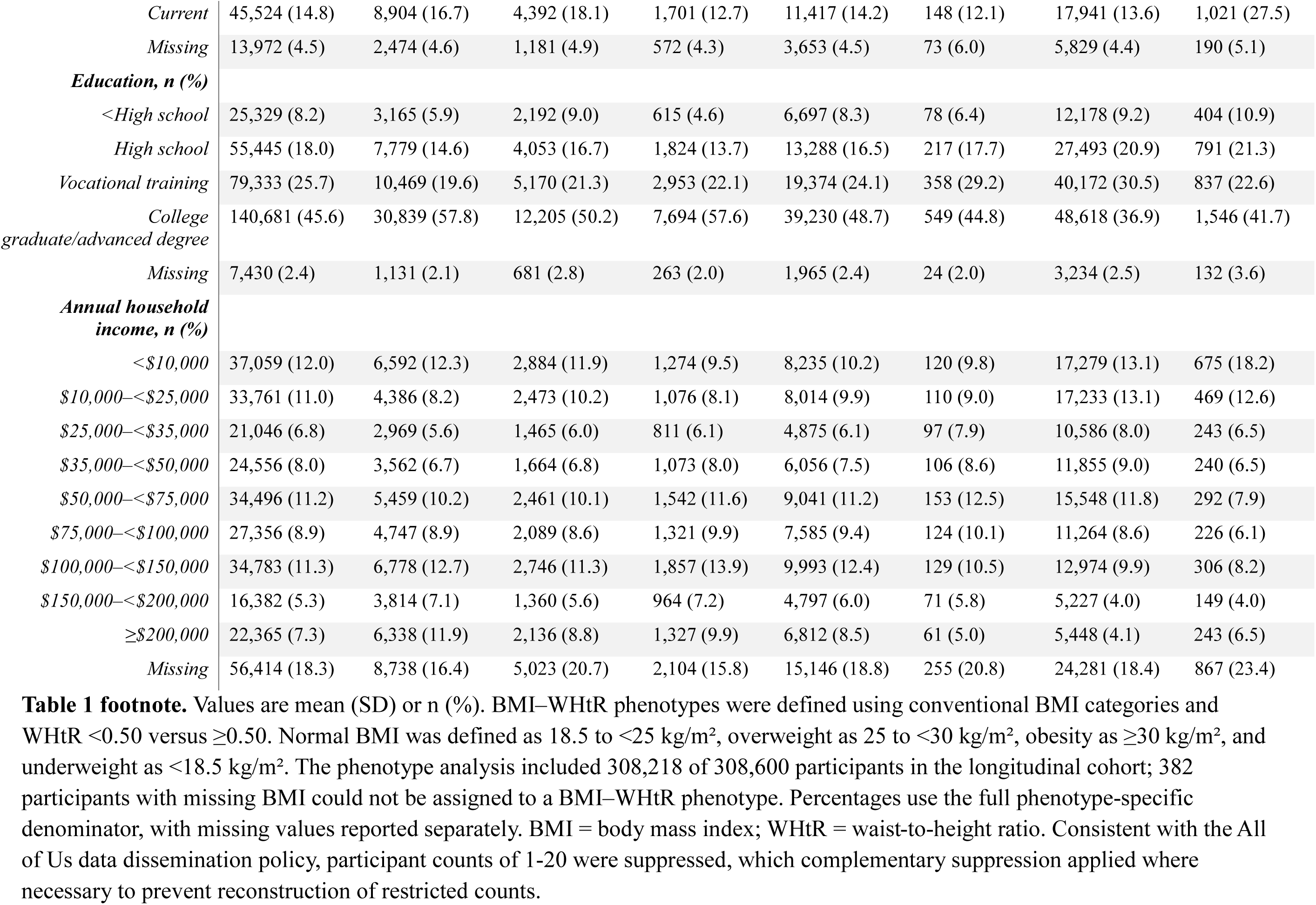
Baseline characteristics according to BMI and waist-to-height ratio phenotype.

Outcome-specific disease-free risk sets ranged from 172,005 participants for hypertension to 283,035 for ischemic stroke. Fully adjusted analytic samples ranged from 117,314 participants for hypertension to 191,396 for ischemic stroke. Event counts ranged from 2,205 for myocardial infarction (MI) to 14,922 for hypertension (**Supplemental Table S7**).

Within the fully adjusted, outcome-specific analytic populations, median follow-up ranged from 2.73 years (IQR, 1.17–4.95) for hypertension to 3.30 years (IQR, 1.46–5.30) for myocardial infarction and ischemic stroke. Total follow-up ranged from 354,405 person-years for hypertension to 649,934 person-years for ischemic stroke. Outcome-specific follow-up durations, person-years, analytic sample sizes, and event counts are provided in **Supplemental Table S8**.

Death censoring had minimal impact on event classification. A small number of diagnoses recorded after death were excluded from the outcome-specific analysis. Outcome-specific counts were withheld in accordance with the AoURP Data and Statistics Dissemination Policy.

### Whole-Cohort Associations Between WHtR and Incident Cardiometabolic Disease

Higher WHtR was associated with increased risk of each cardiometabolic outcome after adjustment for age, sex, BMI, race, ethnicity, smoking status, educational attainment, and household income. Per 0.05-unit higher WHtR, the largest association was observed for T2D (hazard ratio [HR], 1.31; 95% CI, 1.29–1.34). Positive associations were also observed for MI (HR, 1.20; 95% CI, 1.15–1.25), ischemic stroke (HR, 1.16; 95% CI, 1.12–1.20), heart failure (HF; HR, 1.16; 95% CI, 1.14–1.19), chronic kidney disease (CKD; HR, 1.16; 95% CI, 1.13–1.18), IHD (HR, 1.14; 95% CI, 1.11–1.17), and hypertension (HR, 1.14; 95% CI, 1.13–1.16). The association with atrial fibrillation (AF) was smaller but remained positive (HR, 1.07; 95% CI, 1.04–1.09) (**Table 2**; **Figure 1**). Corresponding HRs per 1-SD higher WHtR were 1.77 for T2D, 1.33 for hypertension, 1.31 for IHD, 1.47 for MI, 1.36 for ischemic stroke, 1.37 for HF, 1.14 for AF, and 1.36 for CKD (**Table 2**).

**Figure 1.**
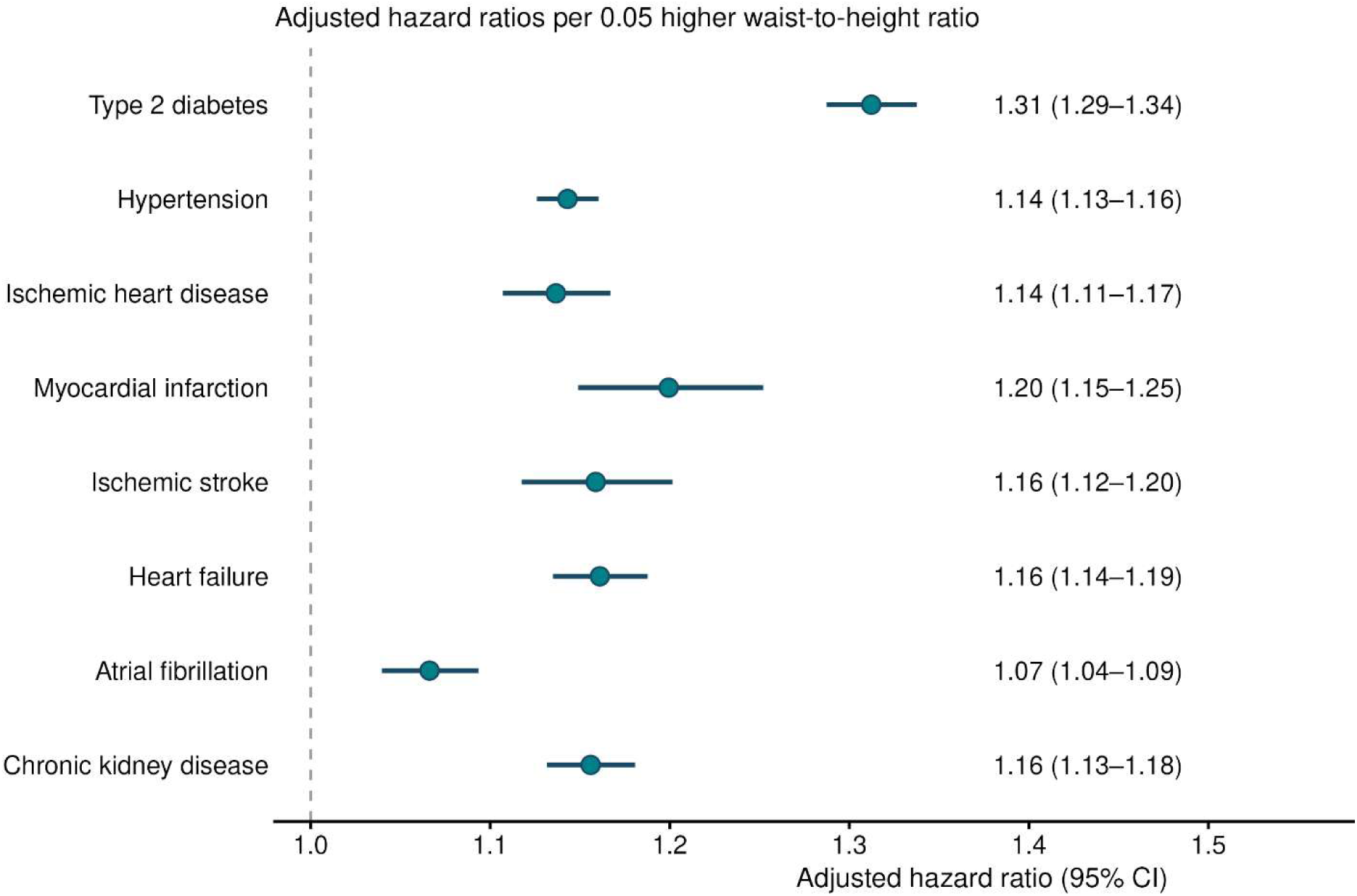
Incident cardiometabolic disease per 0.05-unit higher waist-to-height ratio. Adjusted hazard ratios (HRs) and 95% confidence intervals (CIs) for incident type 2 diabetes, hypertension, ischemic heart disease, myocardial infarction, ischemic stroke, heart failure, atrial fibrillation, and chronic kidney disease per 0.05-unit higher waist-to-height ratio (WHtR). Estimates were obtained from outcome-specific cause-specific Cox proportional hazards models adjusted for age, sex, continuous body mass index (BMI), race, ethnicity, smoking status, educational attainment, and household income. Participants with the corresponding prevalent condition at or before the anthropometric index date were excluded from each outcome-specific analysis.

**Table 2.** Association of waist-to-height ratio with incident cardiometabolic disease in the fully adjusted models.

| ***Outcome*** | ***Analytic N*** | ***Events, N*** | ***Adjusted HR per 0.05 higher WHtR (95% CI)*** | ***P value*** | ***HR per 1-SD higher WHtR (95% CI)*** |
|---|---|---|---|---|---|
| *Type 2 diabetes* | 164,152 | 8,116 | 1.31 (1.29–1.34) | <0.001 | 1.77 (1.72–1.83) |
| *Hypertension* | 117,314 | 14,922 | 1.14 (1.13–1.16) | <0.001 | 1.33 (1.29–1.36) |
| *Ischemic heart disease* | 181,632 | 5,648 | 1.14 (1.11–1.17) | <0.001 | 1.31 (1.25–1.37) |
| *Myocardial infarction* | 188,797 | 2,205 | 1.20 (1.15–1.25) | <0.001 | 1.47 (1.36–1.58) |
| *Ischemic stroke* | 191,396 | 3,073 | 1.16 (1.12–1.20) | <0.001 | 1.36 (1.28–1.45) |
| *Heart failure* | 186,160 | 6,523 | 1.16 (1.14–1.19) | <0.001 | 1.37 (1.32–1.43) |
| *Atrial fibrillation* | 185,933 | 5,593 | 1.07 (1.04–1.09) | <0.001 | 1.14 (1.10–1.19) |
| *Chronic kidney disease* | 182,058 | 8,181 | 1.16 (1.13–1.18) | <0.001 | 1.36 (1.31–1.41) |

**Table 2 footnote.** Hazard ratios (HRs) and 95% confidence intervals (CIs) were estimated using cause-specific Cox proportional hazards models. Follow-up was censored at the earlier of the end of observable follow-up or death. Models were adjusted for age, sex, BMI, race, ethnicity, smoking status, educational attainment, and household income. Events occurring on the recorded date of death were retained; diagnoses occurring after death were excluded. WHtR indicates waist-to-height ratio.

### Nonlinear WHtR Associations in the Whole Cohort

Restricted cubic spline analyses demonstrated heterogeneous exposure-response relationships across outcomes. Evidence of nonlinearity was strongest for T2D, hypertension, HF, and AF; additional evidence was observed for IHD and CKD, whereas evidence was weaker for MI and ischemic stroke. Tests for nonlinearity ranged from *P*<0.001 for several outcomes to *P*=0.041 for ischemic stroke.

T2D and hypertension risks increased nonlinearly at higher WHtR values. IHD, MI, ischemic stroke, and CKD generally showed increasing hazards with higher WHtR. In contrast, HF and AF showed more curvilinear associations, with the lowest estimated hazards near the middle of the WHtR distribution. These findings indicate that WHtR associations cannot be fully characterized by a single linear coefficient or universal dichotomous threshold (**Figure 2; Supplemental Table S9**).

**Figure 2.**
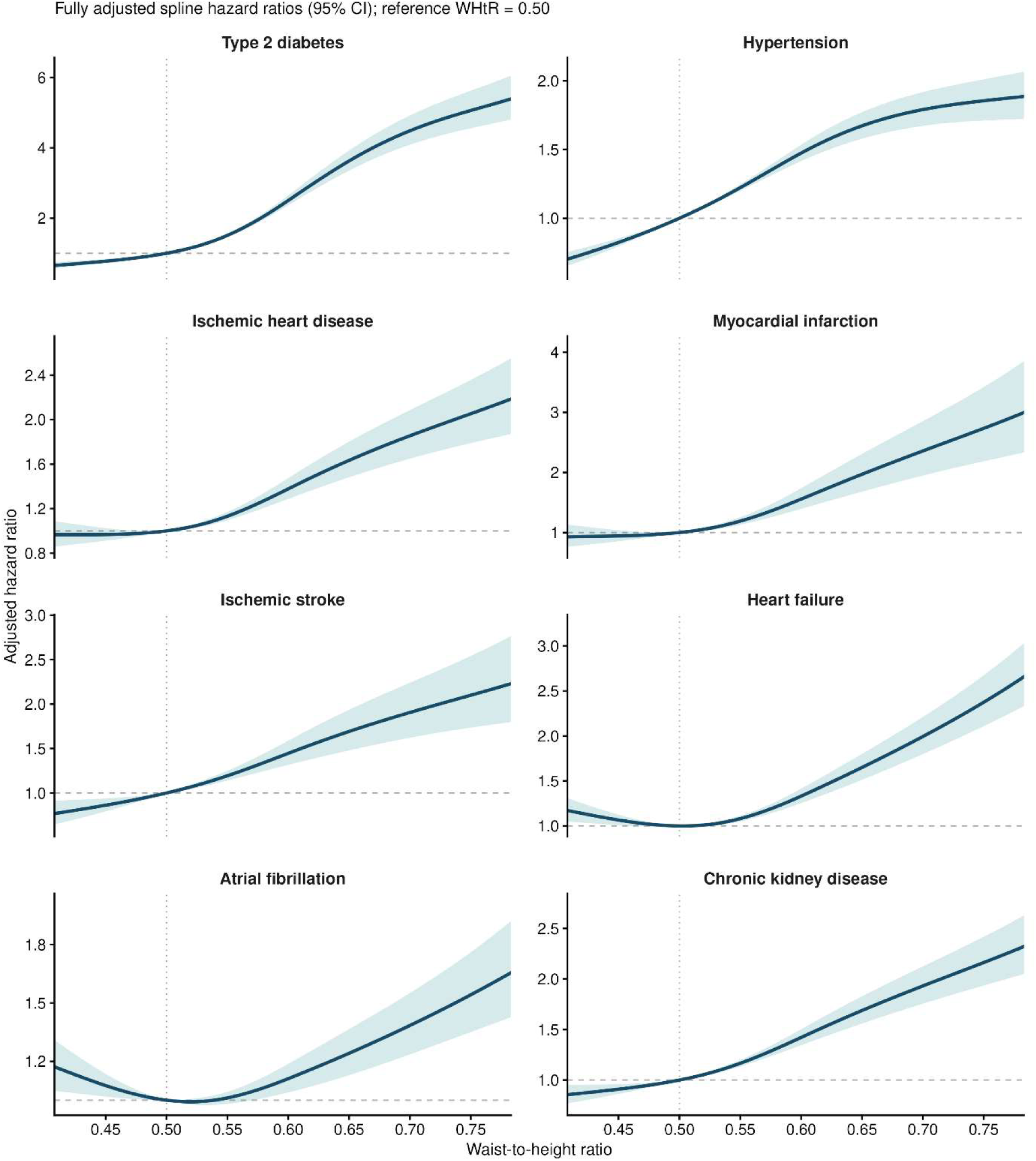
Adjusted cardiometabolic risk across the waist-to-height ratio continuum. Restricted cubic spline associations between WHtR and incident type 2 diabetes, hypertension, ischemic heart disease, myocardial infarction, ischemic stroke, heart failure, atrial fibrillation, and chronic kidney disease. Adjusted HRs are referenced to WHtR 0.50. Solid lines indicate adjusted HRs; shaded bands indicate 95% CIs. Models were adjusted for age, sex, continuous BMI, race, ethnicity, smoking status, educational attainment, and household income. Outcome-specific analyses excluded participants with the corresponding prevalent disease at baseline.

### Cardiometabolic Risk Among Adults With Normal BMI

Among normal-BMI adults, WHtR ≥0.50 versus <0.50 was associated with all eight outcomes after adjustment including continuous BMI (**Table 3**). The largest associations were observed for T2D (HR, 1.74; 95% CI, 1.49–2.03), ischemic stroke (HR, 1.52; 95% CI, 1.27–1.83), and MI (HR, 1.49; 95% CI, 1.19–1.87). Elevated WHtR was also associated with IHD (HR, 1.31; 95% CI, 1.14–1.51), CKD (HR, 1.31; 95% CI, 1.16–1.49), hypertension (HR, 1.26; 95% CI, 1.16–1.37), AF (HR, 1.26; 95% CI, 1.09–1.45), and HF (HR, 1.24; 95% CI, 1.07–1.43) (**Table 3**). Standardized 3-year event probabilities were higher with WHtR ≥0.50 across all outcomes. For WHtR <0.50 versus ≥0.50, probabilities were 1.24% versus 2.14% for T2D, 0.58% versus 0.86% for MI, and 0.80% versus 1.21% for ischemic stroke (**Figure 3**).

**Figure 3.**
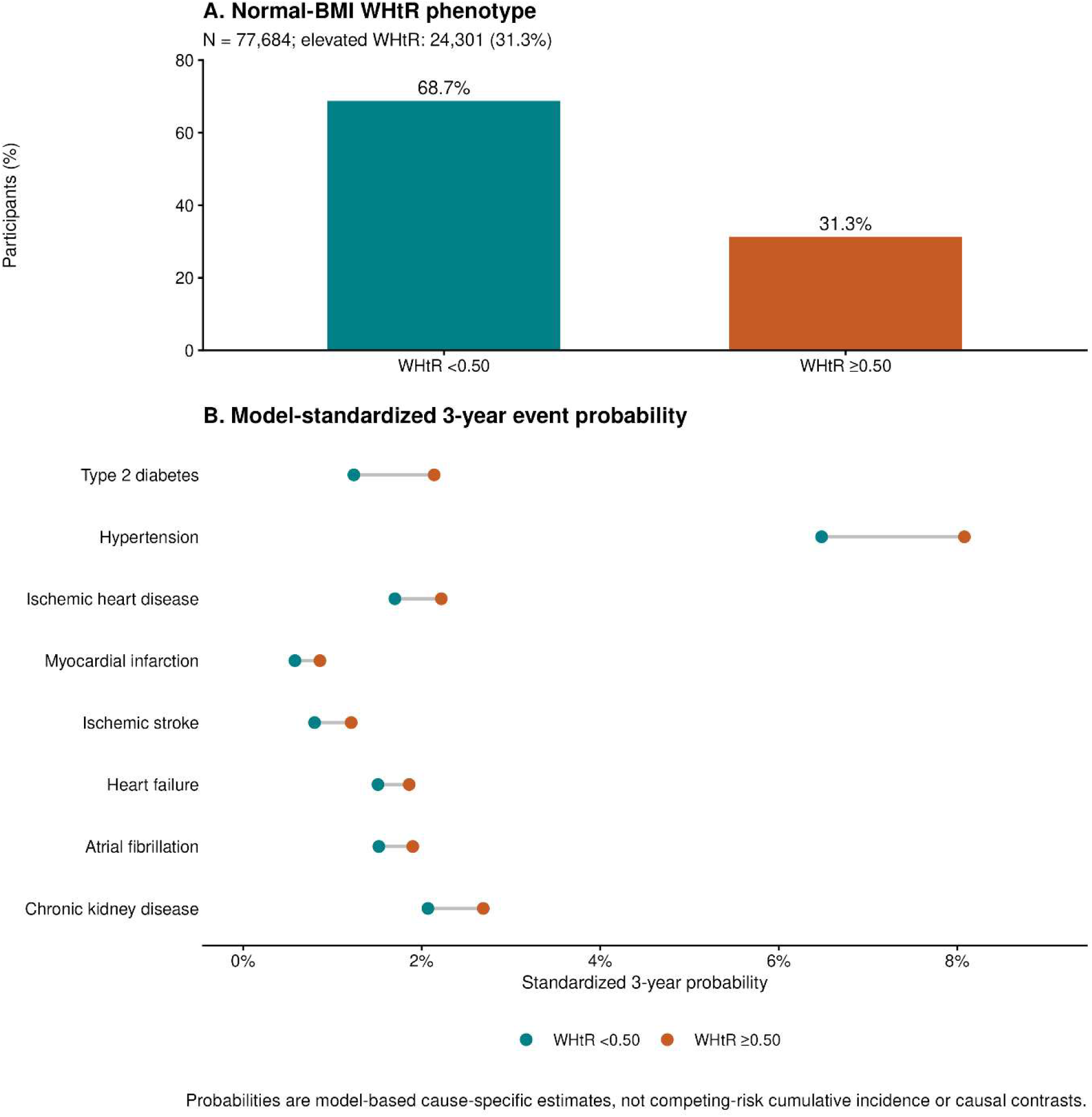
Waist-to-height ratio identifies cardiometabolic risk among adults with normal body mass index. A, Distribution of WHtR categories among 77,684 adults with normal BMI (18.5 to <25 kg/m2). B, Standardized 3-year event probabilities for incident type 2 diabetes, hypertension, ischemic heart disease, myocardial infarction, ischemic stroke, heart failure, atrial fibrillation, and chronic kidney disease according to WHtR <0.50 versus ≥0.50. Estimates were derived from fully adjusted cause-specific Cox models and standardized to the observed distribution of covariates among normal-BMI participants. Models included age, sex, continuous BMI, race, ethnicity, smoking status, educational attainment, and household income. Standardized probabilities are model-based cause-specific event probabilities and should not be interpreted as competing-risk cumulative-incidence estimates or causal effects.

**Table 3.**
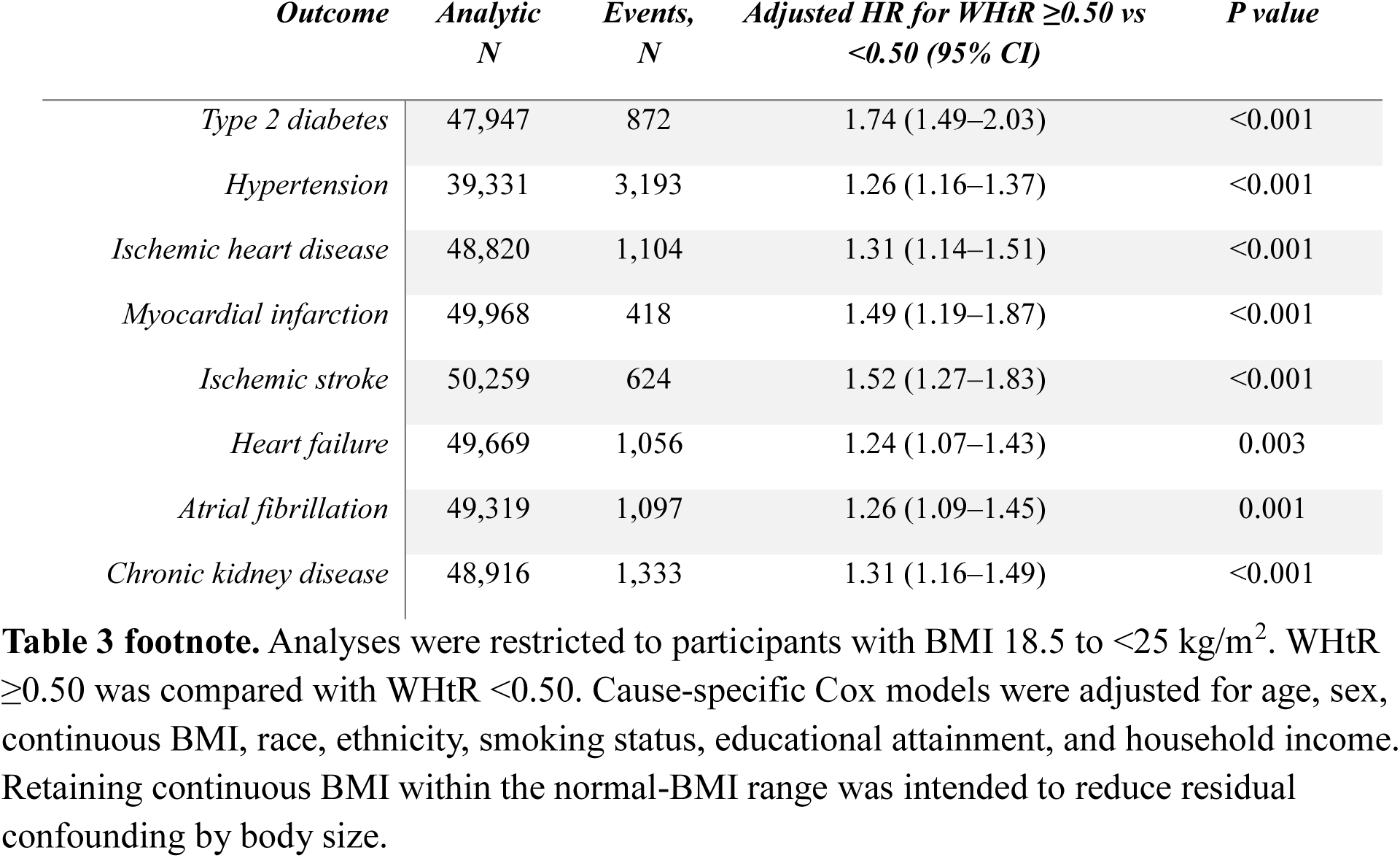
Elevated waist-to-height ratio and incident cardiometabolic disease among adults with normal BMI.

### Comparison of WHtR Associations Across BMI Categories

In additional interaction analyses, the association between continuous WHtR and incident diabetes differed across BMI categories. Adjusted HRs per 0.05-unit higher WHtR were 1.52 (95% CI, 1.43–1.61) among adults with normal BMI, 1.42 (1.36–1.49) among those with overweight, and 1.22 (1.20–1.25) among those with obesity (P for interaction <0.001) (**Supplemental Table S3**). The diabetes association was stronger in normal BMI than obesity (ratio of HRs, 1.24; 95% CI, 1.17–1.32), whereas the comparison with overweight was inconclusive (1.07; 0.99–1.15) (**Supplemental Table S4**).

Hypertension associations also differed across BMI categories. Corresponding HRs were 1.27 (95% CI, 1.22–1.31), 1.18 (1.14–1.21), and 1.08 (1.06–1.10), respectively (P for interaction <0.001). The normal-BMI association was stronger than those in both overweight and obesity (**Supplemental Table S4**). Diabetes and hypertension omnibus interaction tests remained significant after Benjamini–Hochberg adjustment, and findings were similar with flexible age adjustment (**Supplemental Table S3**).

A linear WHtR interaction was also observed for ischemic stroke (P=0.010; Benjamini–Hochberg-adjusted P=0.027), although the flexible-age spline interaction test did not support heterogeneity (P=0.113). Exploratory spline interaction analyses supported heterogeneity for diabetes and hypertension under both age specifications. Additional evidence was observed for myocardial infarction, heart failure, and chronic kidney disease, but these tests do not establish uniformly stronger associations in normal-BMI adults. The linear-age stroke spline interaction model produced a convergence warning and was not interpreted (**Supplemental Tables S3 and S5**).

### Incremental Prognostic Information in the Whole Cohort

Adding nonlinear WHtR improved mean cross-validated discrimination for all eight outcomes. The largest gain was for T2D, with C-index increasing from 0.7343 to 0.7477 (ΔC=0.0134). Gains for other outcomes ranged from 0.0009 for AF to 0.0052 for MI. Linear WHtR provided similar gains for most outcomes, with greater benefits from nonlinear modeling for HF and IHD (**Supplemental Table S6**).

### Sensitivity Analyses

Associations were similar after exclusion of the first 365 days of follow-up. Per 0.05-unit higher WHtR, landmark HRs were 1.31 for T2D, 1.12 for hypertension, 1.13 for IHD, 1.19 for MI, 1.20 for ischemic stroke, 1.15 for HF, 1.06 for AF, and 1.14 for CKD (**Supplemental Table S9**).

Among adults with normal BMI, associations between WHtR ≥0.50 and all eight outcomes persisted in the 365-day landmark analyses. Adjusted HRs were 1.68 (95% CI, 1.39–2.03) for diabetes, 1.20 (1.08–1.32) for hypertension, 1.28 (1.09–1.50) for ischemic heart disease, 1.40 (1.08–1.82) for myocardial infarction, 1.62 (1.31–2.00) for ischemic stroke, 1.21 (1.03–1.43) for heart failure, 1.28 (1.09–1.50) for atrial fibrillation, and 1.19 (1.03–1.38) for chronic kidney disease (**Supplemental Table S10**).

Alternative dichotomous thresholds produced outcome-dependent findings. For T2D, hypertension, IHD, MI, ischemic stroke, and CKD, elevated WHtR generally identified higher risk across thresholds of 0.50, 0.52, and 0.55; HF associations strengthened at higher thresholds, whereas AF showed little or no excess risk at lower thresholds despite a positive continuous association (**Supplemental Figure S2; Supplemental Table S11**).

WHtR-specific proportional-hazards assumptions were generally supported; evidence of nonproportionality was observed for hypertension and AF (**Supplemental Table S12**). For hypertension, the adjusted HR per 0.05-unit higher WHtR was 1.17 (95% CI, 1.14–1.19) during the first 2 years and 1.11 (95% CI, 1.09–1.14) thereafter. Corresponding HRs for AF were 1.08 (95% CI, 1.04–1.12) and 1.05 (95% CI, 1.02–1.09), respectively. Associations therefore attenuated but remained positive during later follow-up (**Supplemental Table S13**).

Multiple-imputation results were highly consistent with complete-case analyses. HRs per 1-SD higher WHtR were 1.75 for T2D, 1.32 for hypertension, 1.29 for IHD, 1.47 for MI, 1.37 for ischemic stroke, 1.41 for HF, 1.15 for AF, and 1.36 for CKD (**Supplemental Table S14**). Associations were robust to flexible adjustment for age. When age was modeled using a natural cubic spline with 4 degrees of freedom, adjusted HRs per 0.05-unit higher WHtR ranged from 1.07 (95% CI, 1.04–1.09) for atrial fibrillation to 1.31 (95% CI, 1.29–1.34) for type 2 diabetes, closely reproducing the primary estimates. Among adults with normal BMI, flexible age adjustment similarly preserved the associations between WHtR ≥0.50 and all eight outcomes, including type 2 diabetes (HR, 1.75; 95% CI, 1.50–2.05), myocardial infarction (HR, 1.50; 95% CI, 1.20–1.88), and ischemic stroke (HR, 1.52; 95% CI, 1.26–1.83). These findings indicate that the principal associations were not materially altered by allowing a nonlinear relationship between age and disease risk (**Supplemental Table S15**).

Sex-stratified analyses demonstrated associations between higher continuous WHtR and all eight outcomes in both females and males. Evidence of a sex interaction was observed for type 2 diabetes (P for interaction=0.003), although the sex-specific estimates were similar in magnitude (HR per 0.05-unit higher WHtR, 1.31 [95% CI, 1.28–1.34] in females and 1.29 [95% CI, 1.25–1.34] in males). No other continuous WHtR association showed a statistically significant sex interaction. Among adults with normal BMI, there was no statistically significant sex interaction for the WHtR ≥0.50 contrast for any outcome. Exploratory omnibus tests identified race-by-WHtR interactions for type 2 diabetes, hypertension, heart failure, and chronic kidney disease, whereas no statistically significant ethnicity-by-WHtR interactions were detected. These exploratory findings require further evaluation and should not be interpreted as evidence of intrinsic biological differences between demographic groups (**Supplemental Tables S15 and S16**).

## Discussion

WHtR identified cardiometabolic risk beyond BMI classification, including among adults with normal BMI. Associations extended across all eight outcomes, with the strongest findings for T2D. The results support complementary assessment of body mass and central adiposity.

Normal BMI did not imply low adiposity-related risk (Sun et al., 2019). Despite a modest BMI difference, the normal-BMI WHtR groups differed by 13.1 cm in waist circumference. Elevated WHtR was associated with a 74% higher T2D hazard and approximately 50% higher MI and stroke hazards, accompanied by higher standardized event probabilities. Previous studies demonstrated within-BMI heterogeneity using waist circumference or waist-to-hip ratio (Jayedi et al., 2022; Feng et al., 2024; Xue et al., 2021). Our contribution extends this evidence across prospective CKM outcomes, standardized event probabilities, and cross-validated discrimination.

BMI-category comparisons strengthen this interpretation. Diabetes associations were stronger in normal BMI than obesity, although the comparison with overweight was inconclusive; hypertension associations were stronger than in both groups. Findings persisted with flexible age adjustment and spline modeling (**Supplemental Tables S3, S4,** and **S5**). These relative associations do not establish greater absolute risk or intervention benefit in normal-BMI adults.

Normal BMI with elevated WHtR might reflect greater abdominal adiposity relative to peripheral lean tissue. Lower muscle mass and insulin resistance could contribute to diabetes risk, but muscle mass, strength, insulin resistance, and diabetes mediation were not assessed. These findings do not establish sarcopenia or a specific mechanism.

Flexible age adjustment was reassuring because elevated-WHtR normal-BMI adults were substantially older. Associations also occurred in both sexes, with similar diabetes estimates despite a formal sex interaction and no sex interaction for the normal-BMI threshold contrast. Exploratory racial differences could reflect body composition, social/environmental exposures, health care access, ascertainment, or residual confounding; they do not establish biological differences or justify race-specific WHtR thresholds. Spline findings support treating WHtR as a continuum. Risk did not change abruptly at 0.50, and HF and AF relationships were more curvilinear than those for T2D and hypertension. Association does not establish prediction or clinical utility. Cross-validated discrimination gains ranged from ΔC=0.0134 for T2D to 0.0009 for AF. The smaller cardiovascular gains suggest limited additional rank-order discrimination beyond the included covariates (Wormser et al., 2011). WHtR appears useful as a complementary phenotype rather than a stand-alone risk model.

Several limitations apply. The observational design precludes causal inference, and residual confounding by lifestyle, medications, family history, body composition, or health care utilization remains possible. Blood pressure, glycemia, and lipids were omitted as potential mediators; their exclusion does not diminish their clinical predictive importance. All of Us is a volunteer cohort, so the 31.3% prevalence is not a national estimate (AoURP Investigators, 2019). Threshold transportability across populations remains uncertain because BMI–adiposity relationships vary (Deurenberg-Yap et al., 2000). Exploratory subgroup analyses involved multiple comparisons and cannot establish subgroup-specific thresholds. Race and ethnicity are social classifications, and these analyses cannot separate contextual exposures from body composition or confounding. EHR outcomes depend on coding, utilization, and observation continuity. Pre-index observation and landmark restrictions reduce but do not eliminate unrecognized prevalent disease, near-term ascertainment, or prior disease-related changes in body composition (**Supplemental Table S10**). BMI interactions were exploratory, and differing WHtR distributions complicate nonlinear comparisons. Baseline-only anthropometry cannot capture subsequent changes. Complete-case analyses retained approximately 67%–69% of disease-free risk sets, although imputation results were concordant. Standardized probabilities are cause-specific model estimates, not competing-risk cumulative incidence. Internal cross-validation does not establish external transportability, calibration, net benefit, or incremental value beyond validated disease-specific risk models.

## Conclusion

WHtR identified prospective cardiometabolic risk that BMI classification alone did not capture. Nearly one-third of adults with normal BMI had WHtR ≥0.50, which was associated with higher incidence of all eight evaluated outcomes, including after exclusion of the first year. Continuous WHtR associations with diabetes and hypertension were particularly pronounced among normal-BMI adults, although heterogeneity differed by outcome and model specification. These findings support assessment of central adiposity alongside BMI.

## Data Availability

The data used in this study are from the All of Us Research Program Registered Tier Curated Data Repository, version 9 (R2025Q4R6; data cutoff January 1, 2025). Individual-level participant data cannot be publicly shared because access is governed by the All of Us Research Program's controlled-access procedures and participant privacy protections. Qualified researchers may apply for access through the All of Us Researcher Workbench, subject to program eligibility requirements, data-use policies, and applicable institutional approvals. Analytic code cannot be publicly released because it was developed and executed within the controlled All of Us Researcher Workbench environment.

## Acknowledgements

The authors thank Professor Shankar Srinivasan of Rutgers University for valuable discussions and continued thoughtful feedback. The authors also gratefully acknowledge the participants of the All of Us Research Program and the All of Us Research Program investigators and staff whose contributions made this research possible.

## Supplemental Material

Figure S1-S2

Tables S1–S16

## Abbreviations

AF: atrial fibrillation
AoURP: All of Us Research Program
BMI: body mass index
CI: confidence interval
CKD: chronic kidney disease
EHR: electronic health record
HF: heart failure
HR: hazard ratio
IHD: ischemic heart disease
MI: myocardial infarction
RCS: restricted cubic spline
T2D: type 2 diabetes
WHtR: waist-to-height ratio

